# Findings from Cardiovascular Evaluation of NCAA Division I Collegiate Student-Athletes after Asymptomatic or Mildly Symptomatic SARS-CoV-2 Infection

**DOI:** 10.1101/2021.01.07.21249407

**Authors:** Calvin E Hwang, Andrea Kussman, Jeffrey W Christle, Victor Froelicher, Matthew T Wheeler, Kegan J Moneghetti

## Abstract

**Objectives:** The risk of myocardial damage after severe acute respiratory syndrome coronavirus 2 (SARS-CoV-2) infection has been controversial. There is an urgent need for data to support the appropriate level of cardiovascular screening for safe return-to-play. The purpose of this study is to report the incidence of abnormal cardiovascular findings in National Collegiate Athletic Association (NCAA) Division I student-athletes with a history of SARS-CoV-2 infection.

**Methods:** This is a case series of student-athletes at a single NCAA Division I institution who tested positive for SARS-CoV-2 by polymerase chain reaction (PCR) or antibody testing (IgG) from 4/15/2020 to 10/31/2020. From 452 athletes who were screened, 5,124 PCR and 452 antibody tests were completed. Student-athletes were followed through 12/31/2020 (median 104 days, range 64-182 days). Cardiac work-up included clinical evaluation, troponin level, electrocardiogram (ECG), and echocardiogram. Additional work-up was ordered as clinically indicated.

**Results:** 55 student-athletes tested positive for SARS-CoV-2. Of these, 38 (69%) had symptoms of Coronavirus Disease (COVID-19), 14 (26%) had a positive IgG test, and 41 (74%) had a positive PCR test. Eight abnormal cardiovascular screening evaluations necessitated further testing including cardiac magnetic resonance imaging (cMRI). Two athletes received new cardiac diagnoses, one probable early cardiomyopathy and one pericarditis, while the remaining six had normal cardiac MRIs.

**Conclusion:** These data support recent publications which recommend the de-escalation of cardiovascular testing for athletes who have recovered from asymptomatic or mildly symptomatic SARS-CoV-2 infection. Continued follow-up of these athletes for sequelae of SARS-CoV-2 is critical.

## INTRODUCTION

With the return of athletic competitions during the SARS-CoV-2 pandemic, the repercussions of infection are still being investigated. Several early studies suggested high rates of cardiac involvement including one cardiac MRI study showing rates of longer term cardiac injury as high as 78%.[1] In the collegiate student-athlete population, a recent cohort study by Rajpal et al. of 26 competitive collegiate athletes with asymptomatic SARS-CoV-2 or mild COVID-19 infections and normal echocardiograms found that 4/26 (15%) had cardiac MRI findings consistent with myocarditis based on the updated Lake Louise criteria.[2] Another cohort study of collegiate student-athletes by Brito et al. found that 56.3% had abnormal findings on echocardiogram and cardiac MRI.[3] However, a study of 18 professional soccer players with a previous history of asymptomatic or mildly symptomatic SARS-CoV-2 infection yielded no signs of cardiac involvement after resting and stress-test ECGs, holter monitoring, echocardiogram, chest CT, and cardiac MRI.[4]

One published expert opinion on cardiac screening for return-to-play after symptomatic COVID-19 infection recommended resting ECG, troponin assay, and 2-D echocardiogram.[5] Consensus opinions were subsequently revised for the collegiate athlete population, with the recommendation for routine cardiovascular testing after mildly symptomatic infections removed.[6] Multiple other groups have published expert recommendations, but there have been few studies to date validating any of these approaches.[7–15] We present a cohort study of 55 collegiate student-athletes with a history of asymptomatic or mildly symptomatic[5] SARS-CoV-2 infection and their subsequent cardiac work-up. The goal of the present study was to determine if current published recommendations were appropriate for detecting post-infection cardiac sequelae.

## METHODS

### Detection of SARS-CoV-2

This prospective observational study included all varsity student-athletes returning to campus from 6/15/2020 to 10/31/2020. A total of 452 student-athletes were included in the study. Upon arrival to campus, all student-athletes underwent serum immunoglobulin G (IgG) antibody (EuroImmun ELISA)[16] and nasopharyngeal or mid-turbinate testing via PCR (Hologic Panther Fusion)[17] for detection of SARS-CoV-2. Student-athletes continued to be tested via surveillance PCR testing one to three times weekly or immediately upon development of any potential COVID-19 symptoms including fever or chills, shortness of breath, sore throat, loss of smell/taste, headache, congestion, chest pain, and cough. All SARS-CoV-2 PCR+ student-athletes were immediately placed in isolation, and symptoms, temperature, and oxygen saturation were recorded daily. Additionally, any student-athlete identified as a potential close contact of a confirmed PCR+ individual was placed in a 14-day quarantine and received COVID-19 PCR testing on days 5 and 12 post-exposure.

### Cardiac Screening Protocol

All PCR+ or IgG+ student-athletes underwent cardiac screening after completion of a ten-day isolation period or upon return to campus if they had tested PCR+ while at home. Asymptomatic individuals with IgG+/PCR-did not complete an isolation period. The cardiovascular screening protocol consisted of a resting 12-lead ECG, troponin-I measurement (Dimension ExL; Siemens; normal <0.055 ng/mL), two-dimensional echocardiogram with doppler, and sports cardiologist consultation. Further testing such as cardiac MRI, ambulatory rhythm monitoring, and cardiopulmonary exercise testing with exercise ECG was at the discretion of the consulting cardiologist. After student-athletes completed the cardiac screening protocol and were cleared by the team physician, they began a gradual return to sport with close monitoring for exercise limitations or recurrent symptoms.

### Patient and Public Involvement

Patients and public were not involved in the design or conduct of the study and did not participate in the analysis of data.

## RESULTS

### Screening and Surveillance Protocol Outcomes

Among the 452 student-athletes who returned to campus starting in June 2020 (**Figure 1**), there were 14 (3.1%) positive and 438 negative antibody tests. 5124 PCR tests were conducted during the study period; 41 (0.8%) were positive and 5083 were negative. Of the 14 IgG+ student-athletes, two recalled loss of smell/taste, three recalled mild upper respiratory symptoms, and nine had no recollection of any COVID-19 symptoms since February 2020. Of the 41 PCR+ student-athletes, 12 were identified on routine surveillance testing, 21 were tested due to symptoms including 12 student-athletes who were tested while at home away from campus, and eight were tested as part of the contact tracing/quarantine protocol. Eight of the 41 (19.5%) were asymptomatic. No student-athlete had moderate symptoms[6] or required hospitalization.

**Figure 1.**
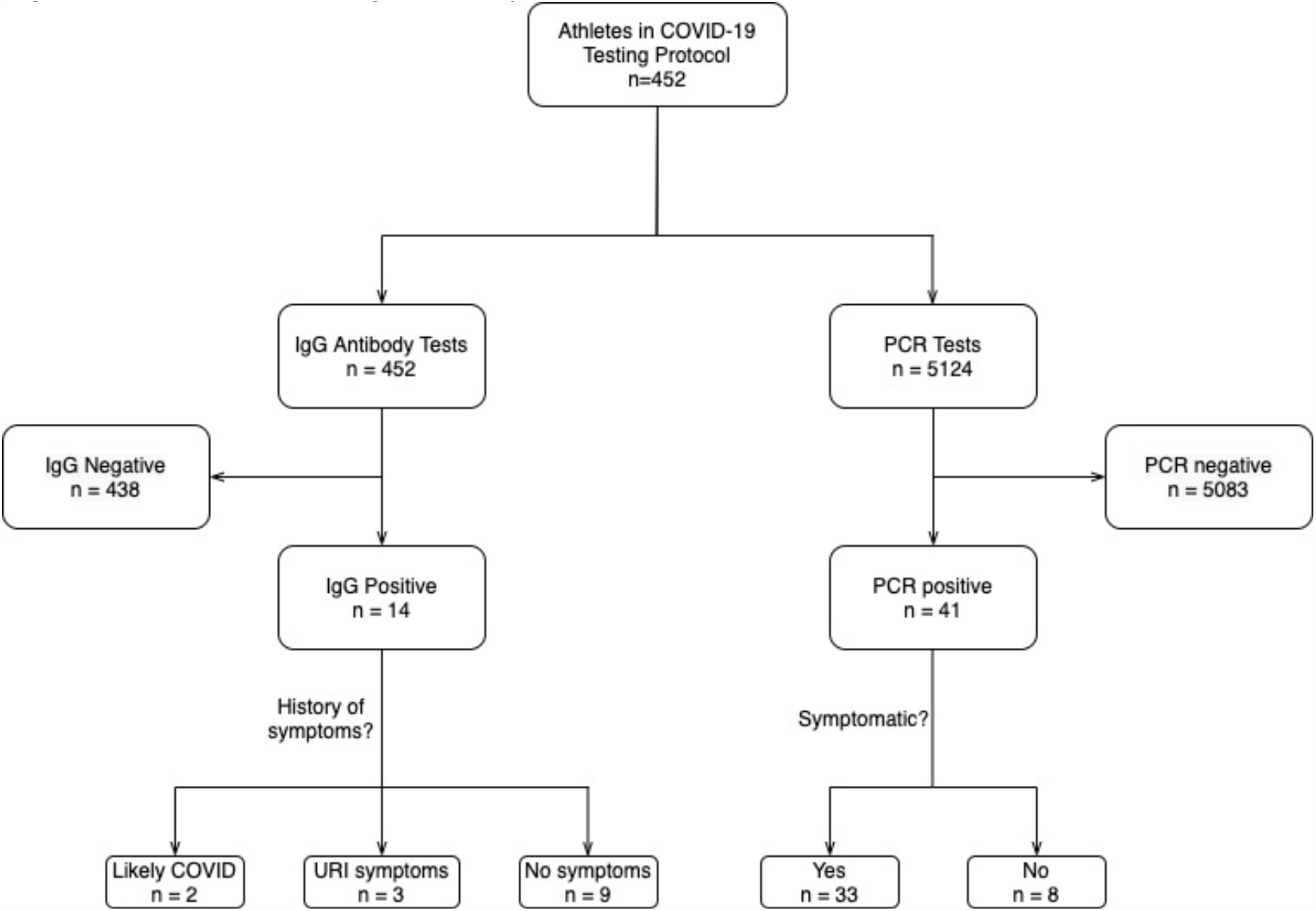
COVID-19 Testing Summary.

### Post-Infection Cardiac Screening

Two of the 14 IgG+ student-athletes had an abnormal initial cardiac work-up, both consisting of a normal ECG/troponin and an abnormal echocardiogram (**Figure 2**); one received an abnormal cardiac diagnosis. This particular student-athlete’s echocardiogram showed a mild reduction in LV and moderate reduction in RV function (no prior echo available). There was no evidence of myocarditis on cardiac MRI. Cardiac MRI confirmed mild biventricular dysfunction as seen on echo. There was no evidence of change in ventricular function on serial imaging over 3 months of follow-up. After further testing and sports cardiology consultation, early cardiomyopathy was diagnosed, thought to be unrelated to his SARS-CoV-2 infection.

**Figure 2.**
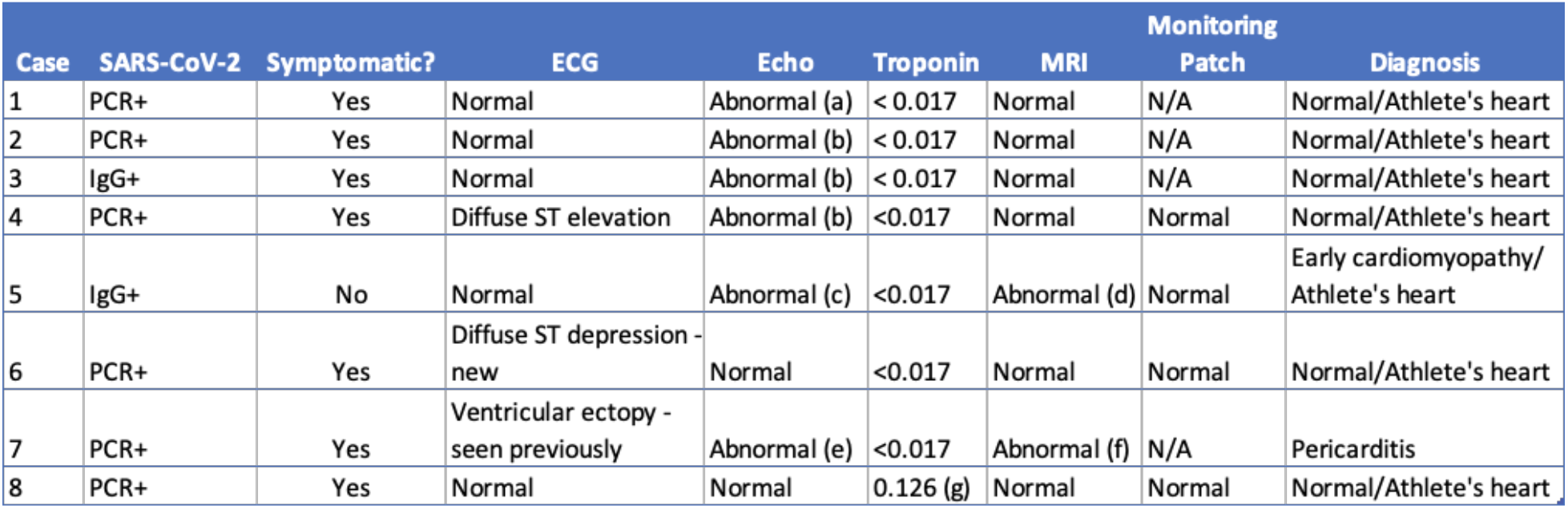
Summary of abnormal cardiovascular screening evaluations a) Borderline LVEF, reduced compared to previous; b) Borderline LVEF; c) Mildly reduced LV function, moderately reduced RV function; d) Mildly dilated LV with LVEF 48% with global hypokinesis without late gadolinium enhancement or abnormal T2 signal. Moderately dilated right ventricle with RVEF 42% and global hypokinesis; e) Mildly reduced LV systolic function, trace to small pericardial effusion. Had normal echo 2 months prior to COVID diagnosis; f) Mild to moderate pericardial effusion, no evidence of myocarditis; g) Repeat troponin level two weeks later was <0.017.

Six out of the 41 PCR+ student-athletes had an abnormal initial cardiac work-up (**Figure 2**), consisting of one with an isolated positive troponin, three abnormal ECGs with abnormal echos, and three normal ECGs with abnormal echos (**Figure 2**). Only one PCR+ student-athlete, who had mild symptoms of sore throat, cough, and congestion at the time of his infection, received an abnormal cardiac diagnosis of pericarditis. He subsequently admitted to having chest pain on exertion prior to receiving clearance to exercise. Interestingly, two months prior to contracting SARS-CoV-2, this student-athlete had ventricular ectopy on resting ECG. At that time, resting echocardiogram was normal. Post-COVID-19 cardiac work-up redemonstrated ventricular ectopy on ECG, but there was a new small pericardial effusion on echocardiogram and cMRI. There was no evidence of myocarditis on cMRI.

Of the 55 PCR+ or IgG+ student-athletes, 13 (23.6%) had changes on echocardiogram reflective of left ventricular remodeling and athletic heart syndrome (AHS). Seven of these 13 student-athletes had a prior echocardiogram which allowed for direct comparison to determine if additional cardiac work-up was indicated. All student-athletes have returned to exercise and athletic activity without complication as of December 31, 2020.

## DISCUSSION

Our study presents the largest cohort to date of collegiate athletes undergoing a post-COVID-19 cardiac screening program. In contrast to studies among community populations, there was a relatively low incidence of cardiac involvement in this young cohort who were asymptomatic or experienced mild symptoms. Second, transiently elevated troponin levels and new abnormal ECG findings did not represent acute myocarditis as assessed by cMRI. Finally, there was a high frequency of left ventricular remodeling representing AHS; this highlights the importance of baseline evaluation in collegiate athletes.

### Evaluation in Asymptomatic or Mildly Symptomatic Cases

Early in the SARS-CoV-2 pandemic, published recommendations included early evaluation of athletes who experience symptomatic infection with ECG, troponin, and echocardiography.[5] Data presented by both Puntmann et al.[1] in unselected patients and Rajpal et al.[2] in competitive athletes highlighted an increased rate of cMRI findings suggestive of myocarditis in those with symptomatic COVID-19. However, in the absence of associated symptoms or abnormal cardiovascular diagnostics to meet clinical criteria for myocarditis,[18] it is unclear if these changes lead to a poor prognosis.[19] These findings present a significant challenge in the development of an athletic screening program for COVID-19 positive athletes. Abnormal findings on cardiac MRI have been reported in up to 50% of asymptomatic athletes,[20] and due to the current prevalence of COVID-19, widespread use of cMRI among athletes is not feasible. Furthermore, the cardiac screening process may result in the detection of abnormalities which do not represent pathology, possibly causing increased stress and delaying return to competition. In order to address these challenges, we developed a screening program which combines severity of disease with detailed cardiovascular system review and implemented a graded approach to the use of cardiovascular diagnostics.

There are very limited data to support or refute the need for comprehensive cardiovascular evaluation in athletes who test positive for SARS-COV-2 but are asymptomatic or have mild symptoms. A report of 30 professional soccer players screened with blood tests, spirometry, ECG, echocardiogram, and stress ECG did not identify clinically relevant anomalies.[4] Data from an Italian professional soccer team suggested a limited role for troponin post-COVID-19 infection.[21] Our data on a relatively large cohort of athletes supports these studies, suggesting a limited role for extensive evaluation in athletes who experience no symptoms or mild symptoms. Within our cohort, one student-athlete had an elevated circulating troponin, proceeded to cMRI, and had no evidence of active myocarditis. Circulating troponin has been shown to be elevated among athletes post-endurance activity,[22–24] and its detection does not always represent cardiac pathology. This individual exercised the morning of the blood draw, which may have led to the abnormal result. Two student-athletes with new ECG findings proceeded to cMRI, which showed no evidence of acute or past myocarditis. These data appear to support more recent expert consensus recommendations,[6] suggesting a limited role for cardiovascular testing following asymptomatic or mildly symptomatic SARS-CoV-2 infection among student-athletes. Continued follow-up of athletes with abnormal cardiovascular studies despite normal cMRI is required to understand the implication of these results.

### Building an Athletic Screening Program

The role of pre-participation cardiac testing with ECG and echocardiogram in young athletes is controversial.[25,26] All matriculated student-athletes in our cohort had a baseline ECG and those on the American football, basketball, and volleyball teams had a baseline echocardiogram. The high incidence of exercise-induced cardiac remodeling added to the complexity in using echocardiography after SARS-CoV-2 infection, particularly as one of the findings of myocarditis is a reduction in LV function.[18] The availability of previous echocardiographic studies in a subset of our student-athletes as part of our pre-participation screening program allowed us to determine if this cardiac remodeling pre-dated their infection, and ultimately reduced the number of cMRIs needed. In the absence of baseline imaging, overreading imaging studies by a team experienced in sports cardiology may help avoid unnecessary investigation or exclusion from training.[27]

### Limitations

Despite being the largest published cohort to date of collegiate student-athletes undergoing post-COVID-19 cardiac screening, our study is still relatively small with limited follow up. Ambulatory rhythm monitoring is recommended in several expert consensus statements, but was not a standard part of our protocol and to our knowledge there is insufficient published data available supporting its routine use in post-COVID-19 cardiac screening. No athlete within our cohort experienced severe COVID-19 symptoms or required hospitalization. The vast majority of student-athletes had previous cardiovascular diagnostics, including 52/55 with a prior ECG and 20/55 with a prior resting echocardiogram available for direct comparison, which helped in interpreting borderline and abnormal findings. The three cases without prior ECGs were first-year student-athletes who had tested positive at home prior to matriculation and had not yet had their pre-participation exam. This cardiovascular screening occurred at a resource-rich institution with an established pre-participation program, and thus these findings may not be practical or representative for all young athletes.

## CONCLUSION

We present data from a relatively large sample of collegiate athletes who tested positive for SARS-CoV-2 during an athletic screening program. In athletes presenting with asymptomatic or mildly symptomatic infections, the rate of cardiac involvement appeared low. The availability of baseline ECGs and echocardiograms was helpful in determining if abnormalities were sequelae of infection. Supportive of recent expert consensus statements, there were no SARS-CoV-2 related cardiac diagnoses in asymptomatic student-athletes. However, larger studies and ongoing follow-up for longer term sequelae of SARS-CoV-2 are vital to the validation of our current conclusions.

### What Are The Findings?

– In contrast to studies performed in community populations post SARS-CoV-2 infection, among a collegiate athletic population who were asymptomatic or experienced mild symptoms, there was a low incidence of cardiac involvement.
– A high frequency of left ventricular remodeling among collegiate athletes, as assessed by echocardiography, highlights the usefulness of baseline evaluation.
– ECG abnormalities and an abnormal troponin did not represent myocarditis as assessed by cardiac MRI.

### How might this affect future practice?

– There appears to be a limited role for extensive cardiovascular diagnostics in collegiate athletes post asymptomatic or mild SARS-CoV-2 infection.
– Larger studies with longitudinal follow-up are required to validate these findings and assess long-term implications of SARS-CoV-2 infection in athletes.

## Data Availability

Data are available upon reasonable request. All data relevant to the study are included in the article or uploaded as supplementary information. Deidentified participant data are available in a database held by Stanford University School of Medicine. They are not available for reuse. Additional information can be requested by contacting the corresponding author.

## Acknowledgments

We would like to acknowledge the athletic trainers and sonographers who contributed to acquisition of the presented data.

## Contributors

All authors gave substantial contribution to the conception or design of the work, or the acquisition, analysis or interpretation of data. All authors gave substantial contribution to drafting the work or revising it critically for important intellectual content. All authors approved the version published. All authors agreed to be accountable for all aspects of the work in ensuring that questions related to the accuracy or integrity of any part of the work are appropriately investigated and resolved. Drs. Hwang and Kussman contributed equally to this work.

## Patient and public involvement

Patients and/or the public were not involved in the design, or conduct, or reporting, or dissemination plans of this research.

## Patient consent for publication

Not required.

## Ethics approval

The study design was reviewed by the Institutional Review Board of Stanford University and determined to be IRB exempt from full review.

